# Sex, Military Occupation, and Rank Are Associated with Risk of Anterior Cruciate Ligament Injury in Tactical-Athletes

**DOI:** 10.1101/2021.09.30.21264383

**Authors:** Aubrey D Aguero, James J Irrgang, Andrew J MacGregor, Scott D Rothenberger, Joseph M Hart, John J Fraser

## Abstract

**Backgroun:** ACL injury is frequent within the U.S. military and represents a significant loss to readiness. Since recent changes to operational tempo, there has not been an analysis of ACL injury risk. There is sparse evidence on salient risk factors for ACL injury across all service members.

**Hypothesis/Purpose:** The aim of this study is to evaluate military occupation, sex, rank, and branch of service on ACL injury risk in the U.S. military from 2006 to 2018.

**Study Design:** Descriptive Epidemiology Study.

**Methods:** The Defense Medical Epidemiology Database was queried for the number of U.S. military members with ICD diagnosis codes 717.83 (Old disruption of ACL), 844.2 (Sprain of knee cruciate ligament), M23.61 (Other spontaneous disruption of ACL), and S83.51 (Sprain of ACL of knee) on their initial encounter from 2006 to 2018. Relative risk (RR) and chi-square statistics were calculated to assess sex and military occupation effects on ACL injury. A multivariable negative binomial regression model evaluated changes in ACL injury incidence with respect to sex, branch of service, and rank.

**Results:** The study period displayed a significant decrease in the ACL injury rate at 0.18 cases per 1000 person-years or relative decrease of 4.08% each year (*p* < 0.001) after averaging over the main and interactive effects of sex, rank, and branch of service. The interaction effect of time with sex indicated a steeper decline in ACL injury incidence in males as compared to females. The risk of ACL injury by sex was modified by rank. Furthermore, the incidence of ACL injury among military personnel varied depending on occupation.

**Conclusion:** Despite the decline in incidence among military members over time, the rates of ACL injury remain much higher than the general U.S. population. Sex, rank, branch of service, and military occupation were found to be risk factors for ACL injury.

**Clinical Relevance:** These results are evidence to support inquiry into the specific hazards associated with these factors. It is critical for policy makers to understand the salient risk factors for ACL injury to guide appropriate proactive measures to prevent injury.

**What is known about this subject:** ACL injury is a known command readiness issue in the military, and there is recent evidence of this within subpopulations of the military.

**What this study adds to existing literature:** This study provides updated trends in ACL injury across the military in light of changes to operational tempo and identifies salient risk factors for ACL injury, which have been previously unknown on a population basis.

## INTRODUCTION

Anterior cruciate ligament (ACL) injury is frequent among athletes^23,25^ and United States military service members^29^ which results in significant costs in terms of time and productivity. The rate of ACL injuries sustained by athletes varies widely by sport and competetive level. In studies of at least moderate sample size, the incidence in professional and elite sports is comparable to that in military populations.^24^ Previous epidemiological studies of ACL injury in the military population have focused on specific subgroups such as the U.S. Naval Special Warfare community^30^ and midshipmen at the U.S. Naval Academy;^18^ however these are both highly active groups that are not representative of the typical service member. In the first military-wide population study assessing ACL injury, Owens and colleagues estimated the incidence of ACL injury for the International Statistical Classification of Diseases and Related Health Problems, Ninth Revision (ICD-9) codes 717.83 (old disruption of ACL) and 844.2 (sprain of knee cruciate ligament) across all services to be 3.0 per 1,000 person-years and 3.7 per 1,000 person-years, respectively, for the period of 1993 to 2003.^27^ This is in stark contrast to the estimated incidence of the general population in the United States at 0.7 per 1,000 person-years.^31^ The burden of ACL injury within the military represents a significant percentage of loss of command readiness^29^ that is further complicated by the changes in operational tempo, the rate of involvment in military activities over time. The end of the study period by Owens and colleagues coincides with the beginning of the conflicts Operation Enduring Freedom (OEF) in 2001 and Operation Iraqi Freedom (OIF) in 2003. The changes to incidence of ACL injury across the U.S. military population since 2003 as a result of the changes in operational tempo remains unclear.

Each military branch is comprised of many, diverse occupations that have specific physical requirements and opportunity for injury. An overview of the diverse military occupational specialties can be viewed at https://bit.ly/MilOcc. Furthermore, rank within the military can be a factor in the amount of physical labor performed. With regards to the risk stratified by rank, most studies have been directed at return to duty rates after ACL reconstruction (ACLR).^5,13^ Antosh and colleagues found that rank was not a significant predictor for return to duty after ACLR at one institution,^5^ however, there is little evidence regarding rank as a risk factor for ACL injury across the U.S. military.

Rescission of the “1994 Direct Ground Combat Definition and Assignment Rule” in January of 2013^14^ allowed for the integration of women into all military occupations employed in combat that were previously closed to them. The Secretary of Defense issued a memorandum to direct all branches to execute their plans to fully implement women into all occupational specialties and career fields no later than April 1, 2016.^11^ Since this time, it is unclear how this policy has affected the incidence of ACL injury by sex within occupational specialties. Analagous to Title IX integrating women in collegiate athletics, observed rates of ACL injuries were higher in male athletes prior to implementation of this policy.^20^ This is despite the documented higher non-modifiable intrinsic risk in women^3,17,23^ and likely a function of exposure. There is an obvious necessity to not only have an updated understanding of the burden of ACL injury to the tactical athlete, but to further elucidate the factors that can increase risk. Therefore, the aim of this epidemiological retrospective cohort study was to evaluate the effects of military occupation, sex, rank, and branch of service on the risk of ACL injury in the U.S. military over the course of time from 2006 to 2018.

## METHODS

A population-based epidemiological retrospective cohort study of all service members in the US Armed Forces was performed to assess the risk of sex, rank, branch of service and military occupation on the incidence of ACL injury. The Defense Medical Epidemiological Database [(DMED), Defense Health Agency, Falls Church, VA, https://bit.ly/DHADMED] was utilized to identify relevant healthcare encounters. This database provides aggregated data for ICD-9 and ICD-10 codes and de-identified patient characteristics, including sex, rank, categories of military occupations, and branch of service for all active duty and reserve military service members. The database is HIPAA compliant, does not include any personal identifiable or personal health information, and has been used previously for epidemiological study of lower extremity injury in the military.^16,27,28^ This study was approved as non–human-subjects research by the Institutional Review Board at the U.S. Naval Health Research Center (NHRC.2020.0203-NHSR).

The database was queried for the number of distinct patients with a primary diagnosis of ACL injury [717.83 (Old disruption of ACL), 844.2 (Sprain of knee cruciate ligament), M23.61 (Other spontaneous disruption of ACL), and S83.51 (Sprain of ACL of knee)] on their initial encounter from 2006 to 2018. Individuals with repeat visits for the same diagnosis were only counted once in all analyses.

Calculations of cumulative incidence of patients diagnosed with an ACL injury were conducted for male and female military members, enlisted and officers, in each service branch (Army, Navy, Marine Corps, and Air Force) and occupational category (enlisted specialties: special operations forces, mechanized/armor, artillery/gunnery, aviation, engineers, maintenance, administration, intelligence, and communication, logistics, and maritime/naval specialties; officer specialties: aviation, engineering and maintenance, administration, operations and intelligence, logistics, and services). Since military end strength fluctuates with a large portion of the force that attrites and is replaced per annum,^22^ the population at risk should be considered a dynamic cohort. Incidence rates were calculated from the sum of the per-annum ACL outcomes and population at risk in the 13-year study epoch. Relative risk (RR) point estimates and 95% confidence intervals (CIs), risk difference point estimates, attributable risk (AR), number needed to harm (NNH), and chi-square statistics were calculated to assess the effects of sex and military occupational category. All of the preceeding calculations were performed using Microsoft Excel for Mac 2016 (Microsoft Corp., Redmond, WA) and a custom epidemiological calculator spreadsheet.^21^

A multivariable negative binomial regression was performed to evaluate time trends with respect to ACL injury incidence and included the factors of sex, rank, and branch of service. Average marginal effects of predictor variables in the negative binomial model were estimated for ease of interpretation. Due to overdispersion of the data indicated by the likelihood ratio test demonstrating that alpha is significantly different from zero, the negative binomial model was chosen over the Poisson regression model.^10^ The regression analysis was performed using Stata 16 software (StataCorp. 2019. Stata Statistical Software: Release 16. College Station, TX: StataCorp LLC.).

Male service members were used as the reference group in the assessment of sex. Enlisted personnel were used as the reference group for rank as there is evidence that enlisted personnel suffer more disease and non-battle injuries as compared to commissioned officers.^8^ Enlisted Infantry and Ground/ Naval Gunfire officer groups were used as the reference groups in the assessment of occupational risk. From an organizational and operational perspective, injury risk is referenced to groups with increased exposures to injury due to the relatively higher physical requirements of these occupational categories.^4,16^ Finally, the Army was the reference group for service branch, with evidence supporting increased injury rates in the Army compared to the other services.^19^ The level of significance was *p* ≤ .05 for all analyses, and no adjustments were made for multiplicity. Relative risk point estimates were considered statistically significant if CIs did not cross the 1.00 threshold.

## RESULTS

During the study period from 2006 to 2018, 59,555 enlisted males sustained ACL injuries (4.8 per 1000 person-years) and 8,350 enlisted females sustained ACL injuries (3.9 per 1000 person-years) for a total of 67, 905 enlisted member ACL injuries across the services (4.6 per 1000 person-years). During the same time period, 9,983 male officers sustained ACL injuries (4.0 per 1000 person-years) and 2,198 female officers sustained ACL injuries (4.5 per 1000 person-years) for a total of 12,181 officer ACL injuries across the services (4.1 per 1000 person-years). Tables 1 and 2 display the counts and incidence rates. The overall trend across all services, sex, and rank was a decreasing rate of ACL injury with the exception of spikes in incidence in 2010 and 2016 for female Marine Corps officers (Figures 1-4). Officer male incidence per 1000 person-years decreased from 5.2 (95% CI, 4.9-5.5) to 2.7 (95% CI, 2.5 to 2.9), officer female incidence per 1000 person-years decreased from 5.1 (95% CI, 4.3-5.9) to 3.7 (95% CI, 3.1-4.3), enlisted male incidence per 1000 person-years decreased from 5.9 (95% CI, 5.7-6.1) to 3.3 (95% CI, 3.2-3.4), and enlisted female incidence per 1000 person-years decreased from 4.5 (95% CI, 4.2-4.8) to 2.8 (95% CI, 2.6-3.0).

**Table 1:** Number and Incidence of ACL Injury among Officers in the US Armed Forces by Sex and Occupation, 2006-2018.

| Officer Specialty | Ground and Naval Gunfire |  |  | Aviation |  |  | Engineering & Maintenance |  |  | Administration |  |  | Operations & Intelligence |  |  | Logistics |  |  | Services |  |  | Total |  |  |
| --- | --- | --- | --- | --- | --- | --- | --- | --- | --- | --- | --- | --- | --- | --- | --- | --- | --- | --- | --- | --- | --- | --- | --- | --- |
| ACL Injuries (n) |  |  |  |  |  |  |  |  |  |  |  |  |  |  |  |  |  |  |  |  |  |  |  |  |
|  | M | F | Total | M | F | Total | M | F | Total | M | F | Total | M | F | Total | M | F | Total | M | F | Total | M | F | Total |
| Army | 1110 | 24 | 1134 | 442 | 28 | 470 | 736 | 121 | 857 | 276 | 116 | 392 | 428 | 120 | 548 | 430 | 122 | 552 | 785 | 364 | 1149 | 4328 | 913 | 5241 |
| Navy | 388 | 49 | 437 | 435 | 37 | 472 | 279 | 23 | 302 | 80 | 23 | 103 | 151 | 32 | 183 | 100 | 12 | 112 | 351 | 200 | 551 | 2017 | 444 | 2461 |
| Air Force | 51 | 2 | 53 | 693 | 42 | 735 | 397 | 72 | 469 | 171 | 81 | 252 | 356 | 124 | 480 | 266 | 87 | 353 | 494 | 285 | 779 | 2673 | 742 | 3415 |
| Marines | 201 | 2 | 203 | 197 | 13 | 210 | 102 | 9 | 111 | 77 | 15 | 92 | 106 | 6 | 112 | 124 | 22 | 146 | 16 | 3 | 19 | 965 | 99 | 1064 |
| Total | 1750 | 77 | 1827 | 1767 | 120 | 1887 | 1514 | 225 | 1739 | 604 | 235 | 839 | 1041 | 282 | 1323 | 920 | 243 | 1163 | 1646 | 852 | 2498 | 9983 | 2198 | 12181 |
| Incidence (per 1000 person-years) |  |  |  |  |  |  |  |  |  |  |  |  |  |  |  |  |  |  |  |  |  |  |  |  |
| Army | 4.5 | 7.1 | 4.5 | 4.0 | 4.5 | 4.1 | 4.6 | 4.6 | 4.6 | 4.0 | 4.4 | 4.1 | 4.6 | 6.7 | 4.9 | 4.7 | 4.7 | 4.7 | 3.9 | 4.3 | 4.0 | 4.3 | 4.7 | 4.4 |
| Navy | 3.4 | 3.5 | 3.4 | 3.5 | 4.5 | 3.5 | 3.8 | 5.2 | 3.9 | 3.0 | 3.2 | 3.0 | 4.0 | 5.0 | 4.1 | 3.6 | 2.5 | 3.4 | 3.3 | 3.6 | 3.4 | 3.5 | 3.9 | 3.6 |
| Air Force | 5.0 | 1.2 | 4.5 | 3.4 | 3.8 | 3.5 | 4.4 | 5.0 | 4.5 | 4.2 | 4.8 | 4.4 | 3.7 | 5.4 | 4.1 | 5.4 | 6.6 | 5.7 | 3.9 | 3.9 | 3.9 | 4.0 | 4.6 | 4.1 |
| Marines | 3.9 | 3.2 | 3.9 | 3.7 | 7.9 | 3.8 | 3.5 | 5.8 | 3.6 | 3.0 | 3.8 | 3.1 | 4.3 | 3.4 | 4.2 | 3.9 | 5.3 | 4.0 | 2.4 | 3.2 | 2.5 | 3.8 | 5.7 | 3.9 |
| Total | 4.1 | 3.9 | 4.1 | 3.6 | 4.4 | 3.7 | 4.3 | 4.8 | 4.3 | 3.7 | 4.3 | 3.9 | 4.1 | 5.8 | 4.4 | 4.6 | 5.0 | 4.7 | 3.7 | 3.9 | 3.8 | 4.0 | 4.5 | 4.1 |
| Occupation Codes | O205: Ground and Naval Arms; O206: Missiles |  |  | O201: Fixed-Wing Fighter/Bomber Pilots; O202: Other Fixed-Wing Pilots; O203: Helicopter Pilots; O204: Aircraft Crews |  |  | OFF4: Engineering and Maintenance Officers |  |  | OFF1: General Officers and Executives, N.E.C.; OFF7: Administrators |  |  | O207: Operations Staff; O301: General Intelligence; O302: Communications Intelligence; O303: Counterintelligence |  |  | OFF8: Supply, Procurement and Allied Officers |  |  | OFF5: Scientists and Professionals; OFF6: Health Care Officers |  |  | All Officer Specialties |  |  |

**Table 2:**
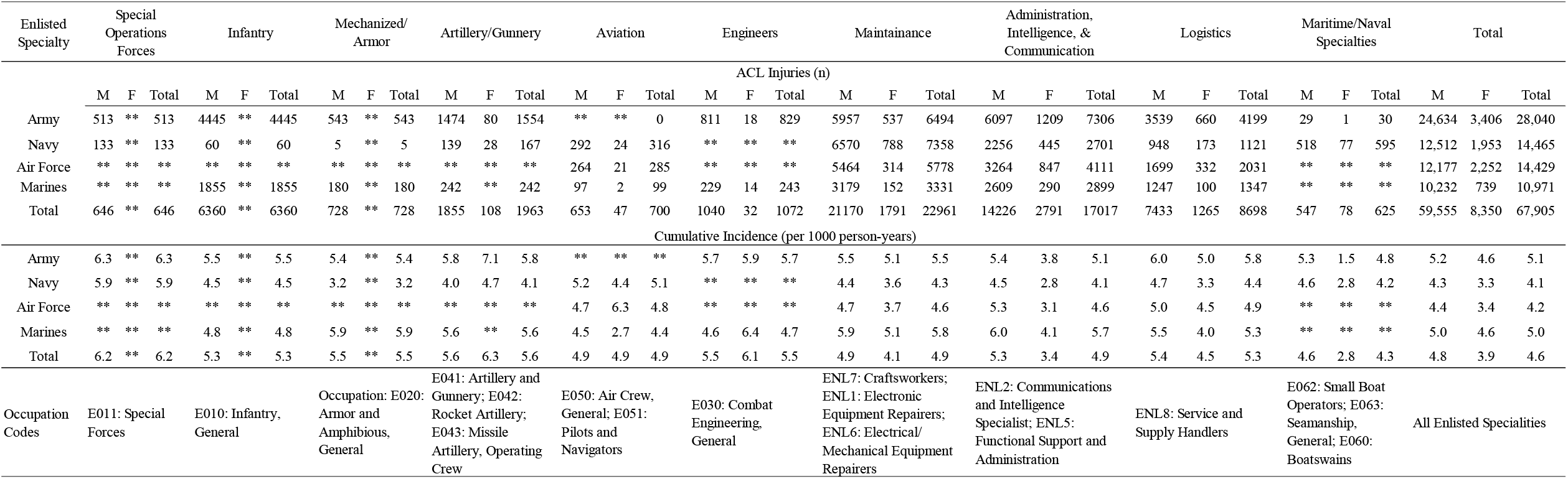
Number and Incidence of ACL Injury among Enlisted Members in the US Armed Forces by Sex and Occupation, 2006-2018.

**Figure 1.**
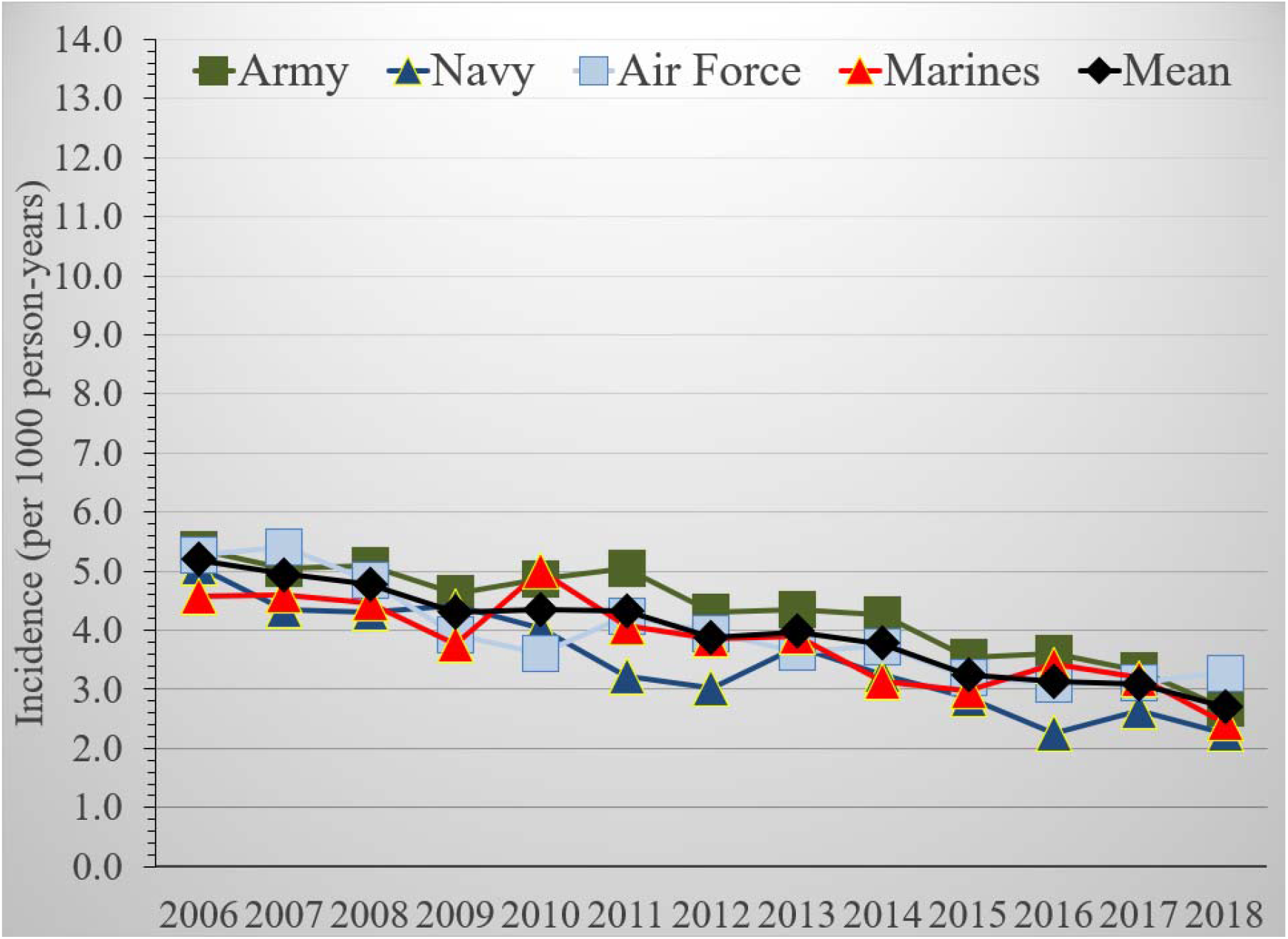
Anterior Cruciate Ligament Injury Incidence among Male Officers, US Armed Forces, 2006–2018.

**Figure 2.**
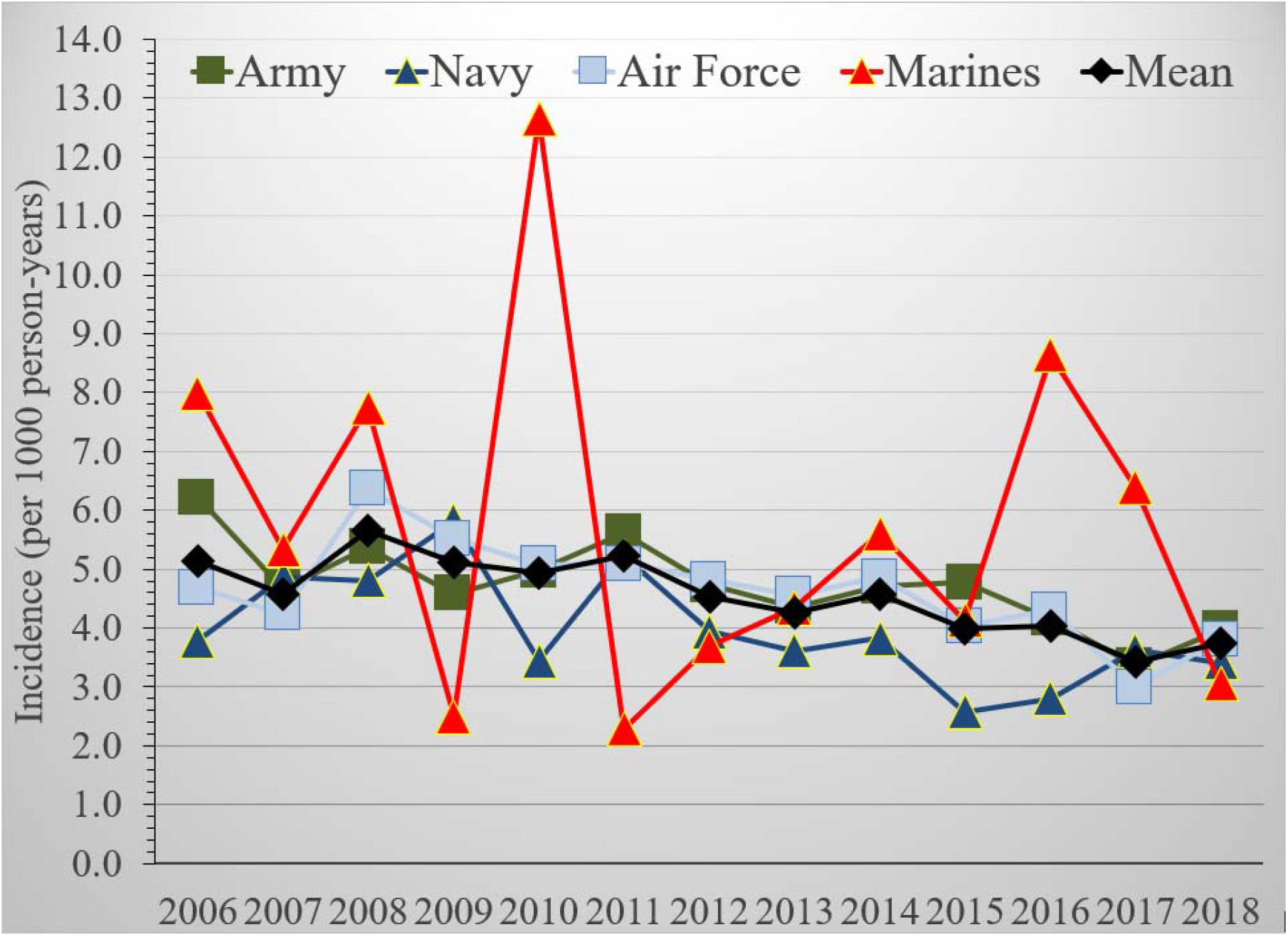
Anterior Cruciate Ligament Injury Incidence among Female Officers, US Armed Forces, 2006–2018.

**Figure 3.**
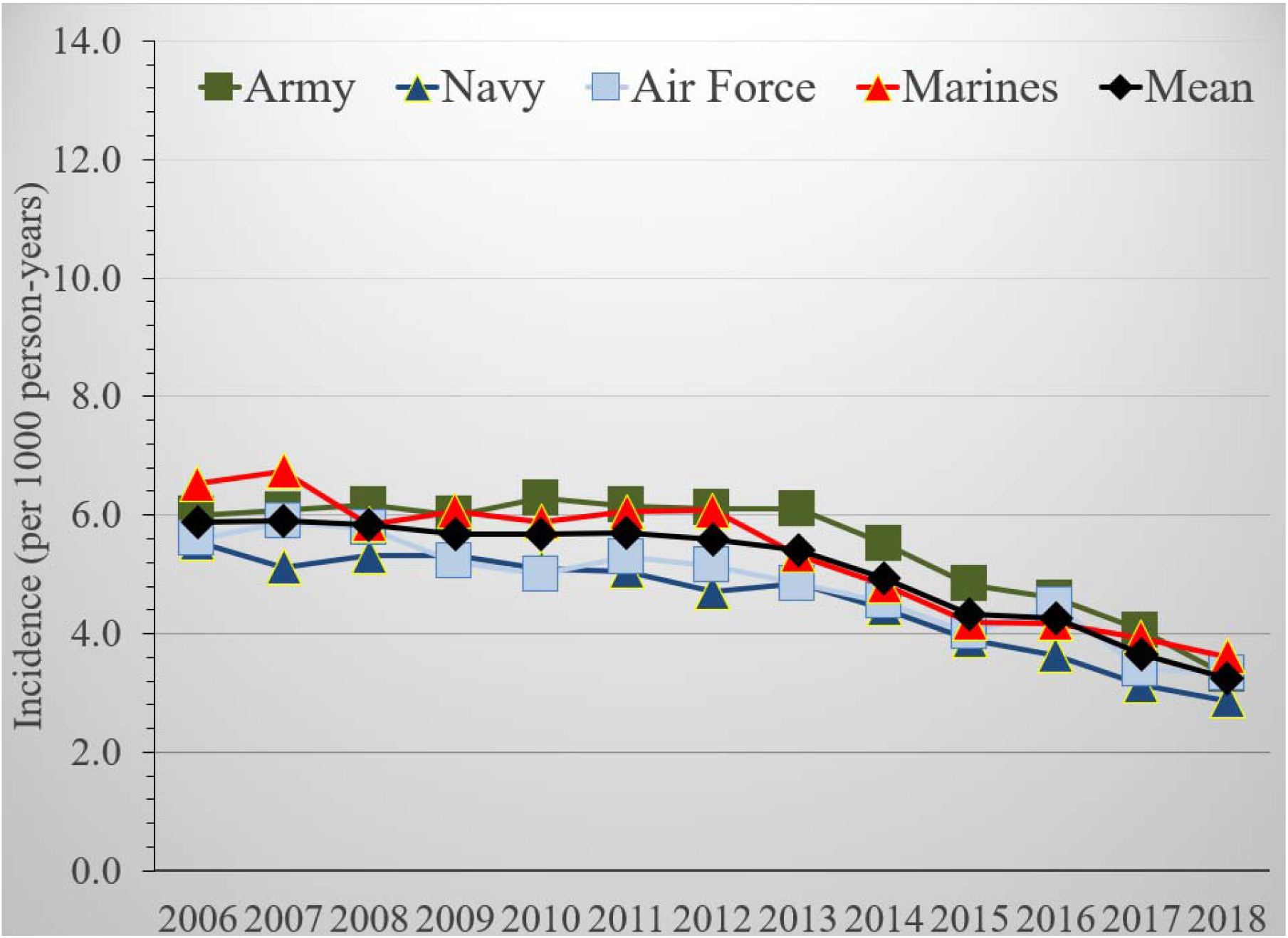
Anterior Cruciate Ligament Injury Incidence among Enlisted Males, US Armed Forces, 2006–2018.

**Figure 4.**
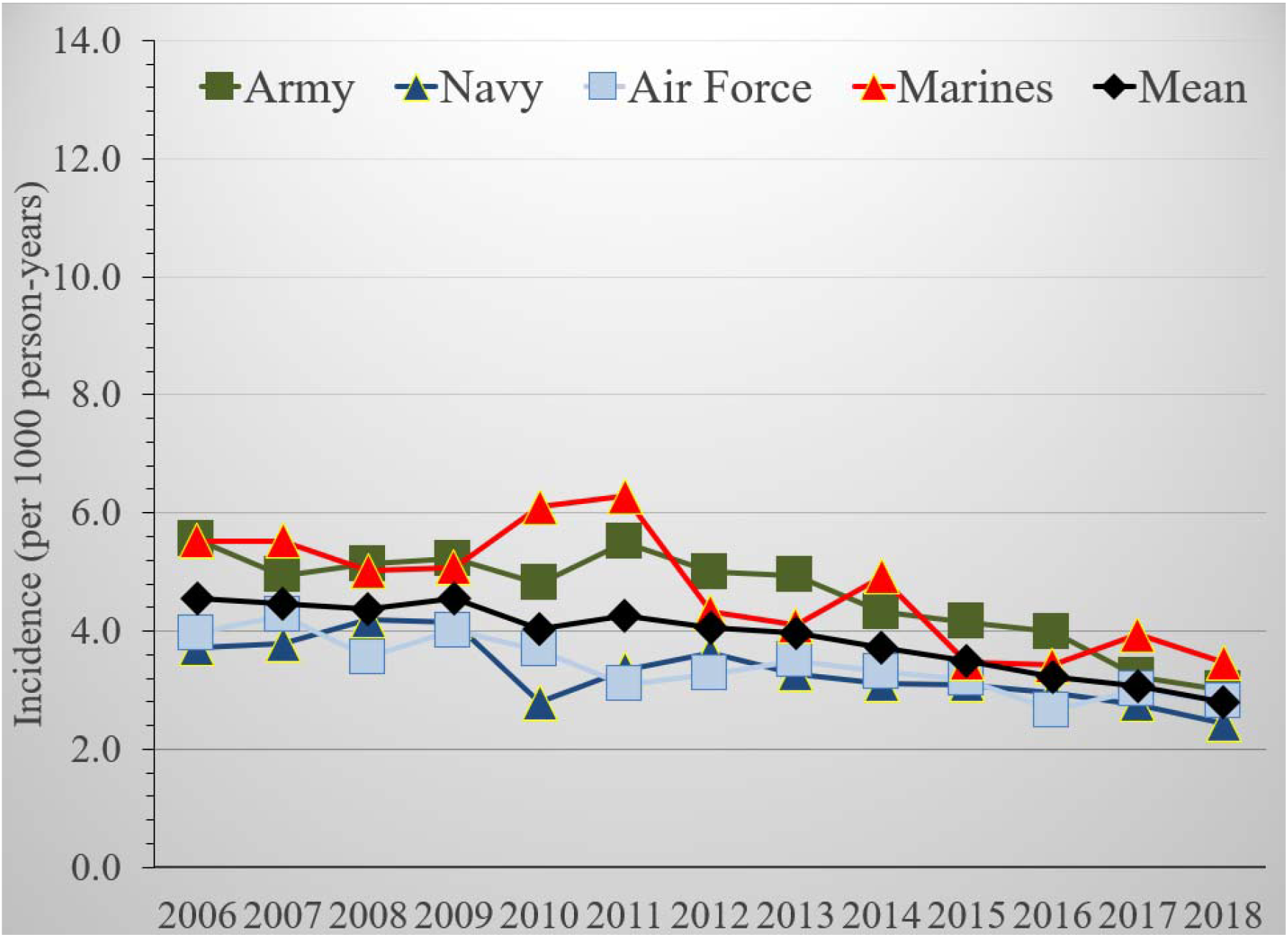
Anterior Cruciate Ligament Injury Incidence among Enlisted Females, US Armed Forces, 2006–2018.

Table 3 reports the risk of ACL injury by sex, with females referenced to males within their respective rank (officer versus enlisted) and occupation. Female officers had a statistically significant increased risk of ACL injury overall compared to their male colleagues (RR = 1.14 (95% CI 1.09-1.19), AR = 12.0%, NNH = 1836, *p* <.001) and in aviation, administration, and operations and intelligence occupations (RR range = 1.16-1.39, AR = 14-28.1%, NNH = 617-1650, *p* ≤ .05). Female enlisted members had a statistically significant decreased risk of ACL injury overall (RR = 0.82 (95% CI = 0.80-0.83), AR = -22.7%, NNH = -1134, *p* <.001) and in maintenance, administration, intelligence, and communication, logistics, and maritime/naval specialties compared to males (RR range = 0.61-0.83, AR = -64 – -20.9%, NNH = -1173 – -523, *p* < .001).

**Table 3:**
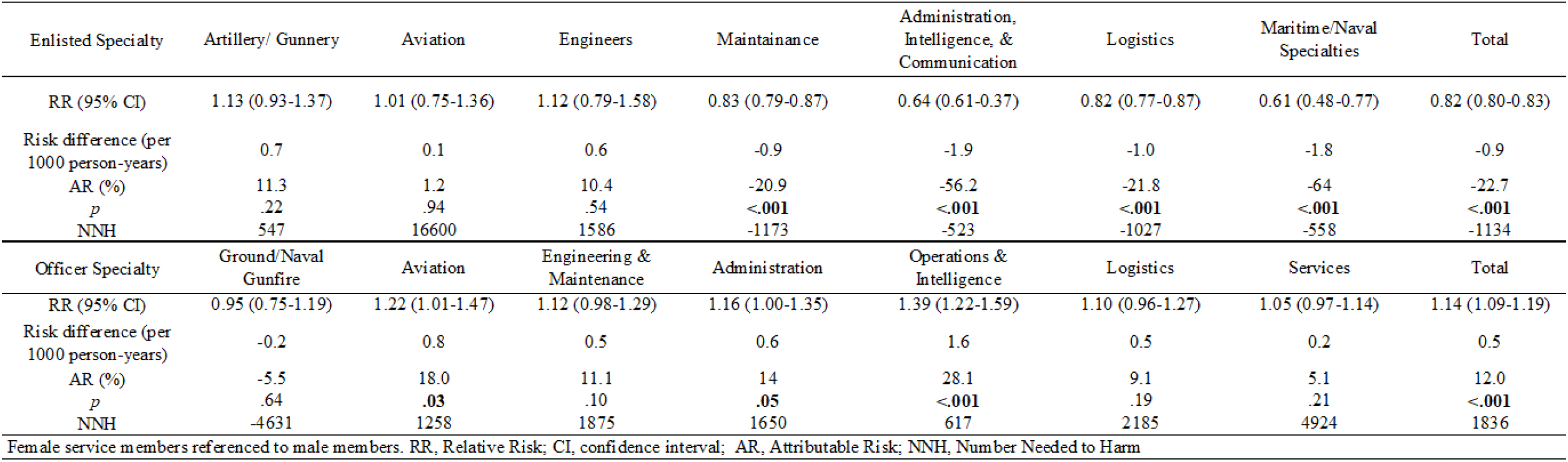
Risk of ACL Injury by Sex in Members of the US Armed Forces, 2006-2018.

Occupation is shown as a risk factor for ACL injury referenced to infantry for enlisted occupations and referenced to ground and naval gunfire officers for officer occupations in Table 4. Enlisted personnel in aviation, maintenance, administration, intelligence, communication, and maritime/naval specialties were at a statistically significant lower risk compared to infantry (RR range = 0.81-0.93, AR = -23.5 – -8.0%, NNH = -2576 – -1002, *p* ≤ .05), and Special Operation Forces and Artillery/Gunnery occupations were at a statistically significant higher risk compared to infantry (RR range = 1.07-1.19, AR = 6.3 – 15.8%, NNH = 1013 – 2827, *p* ≤ .01). Aviation officers and services officers had a statistically significant lower risk of ACL injury compared to ground and naval gunfire officers (RR range = 0.89-0.92, AR = -12.9 – -8.2%, NNH = -2127 – -3187, *p* ≤ .01), and logistics officers had a statistically higher risk compared to ground and naval gunfire officers (RR = 1.13, AR = 11.8%, NNH = 1808, *p* <.001).

**Table 4:**
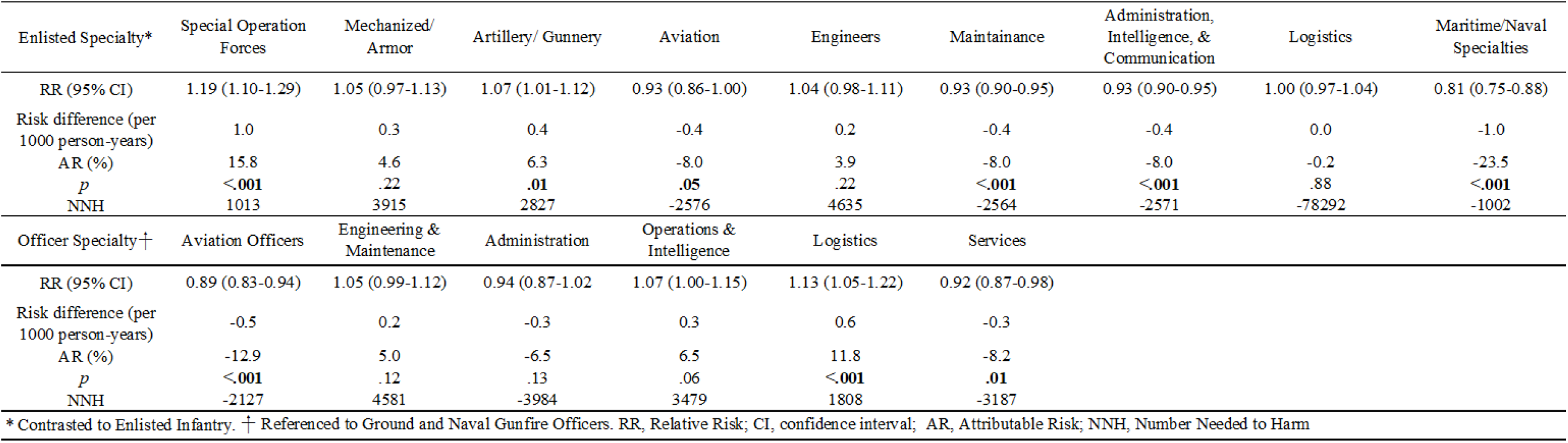
Risk of ACL Injury by Occupation in Members of the US Armed Forces, 2006-2018.

**Table 5:** Negative Binomial Regression Model for Incident Rate Ratio Estimates of ACL Injury over Time, adjusted for Sex, Rank, and Branch of Service, in Members of the US Armed Forces, 2006-2018.

| Variable* | IRR | Standard Error | 95% Confidence Interval | *p* |
| --- | --- | --- | --- | --- |
| Year | 0.954 | 0.00318 | 0.947 – 0.960 | <0.001 |
| Sex | $5.50 \times 10^{-11}$ | $6.13 \times 10^{-10}$ | $1.78 \times 10^{-20} - 0.169$ | 0.03 |
| Sex x Year interaction | 1.012 | 0.00561 | 1.001 – 1.023 | 0.04 |
| Rank | 0.893 | 0.0187 | 0.858 – 0.931 | <0.001 |
| Branch of Service: USN | 0.792 | 0.211 | 0.752 – 0.835 | <0.001 |
| Branch of Service: USAF | 0.866 | 0.0227 | 0.823 – 0.912 | <0.001 |
| Branch of Service: USMC | 0.961 | 0.0281 | 0.908 – 1.018 | 0.17 |

\*The interaction terms of Rank x Year and Branch of Service x Year are not included in final model with $p = 0.84$ and $0.69$ , respectively. Reference for Sex is Male, for Rank is Enlisted, and for Branch of Service is Army.

The results of the multivariable negative binomial regression demonstrated significant effects of time, sex, rank, branch of service, and an interaction effect of sex with time on the incidence of ACL injury. There is a statistically significant decrease in ACL injury incidence by 4.64% (95% CI = 4.02 – 5.27%, *p* < 0.001) per year in the reference group of enlisted males in the US Army. Officers in comparison to enlisted personnel have 0.89 times the rate of ACL injury (95% CI = 0.86 – 0.93, *p* < 0.001). The US Navy and US Air Force have 0.79 times (95% CI = 0.75 – 0.84, *p* < 0.001) and 0.87 times (95% CI = 0.82 – 0.91, *p* < 0.001) the rate of the US Army, respectively, while the US Marine Corps was not statistically different in comparison to the Army (0.96, 95% CI = 0.91 – 1.02, *p* = 0.17). Through calculation of the average marginal effect, the decrease in ACL injury incidence is 0.18 cases per 1000 person-years (95% CI = 0.16 – 0.20 per 1000 person-years, *p* < 0.001), after averaging over the main and interactive effects of sex, rank, and branch of service. This decrease in ACL injury incidence is a 4.08% relative reduction in the injury rate per year (95% CI = 3.56% - 4.60%, *p* < 0.001).

Figure 5 depicts the interaction effect of time and sex on the outcome of ACL injury incidence. Through calculation of the average marginal effect for year, a statistically significant difference was found in the rate of decrease between males and females (*p* = 0.005). The decrease in incidence in males is 0.22 cases per 1000 person-years (95% CI = 0.18 – 0.25, *p* < 0.001) per year compared to females at 0.15 cases per 1000 person-years (95% CI = 0.11 – 0.18, *p <* 0.001) per year, with the difference in the rate of decrease per year at 0.069 cases per 1000 person-years (95% CI = 0.021 – 0.12, *p =* 0.005).

**Figure 5:**
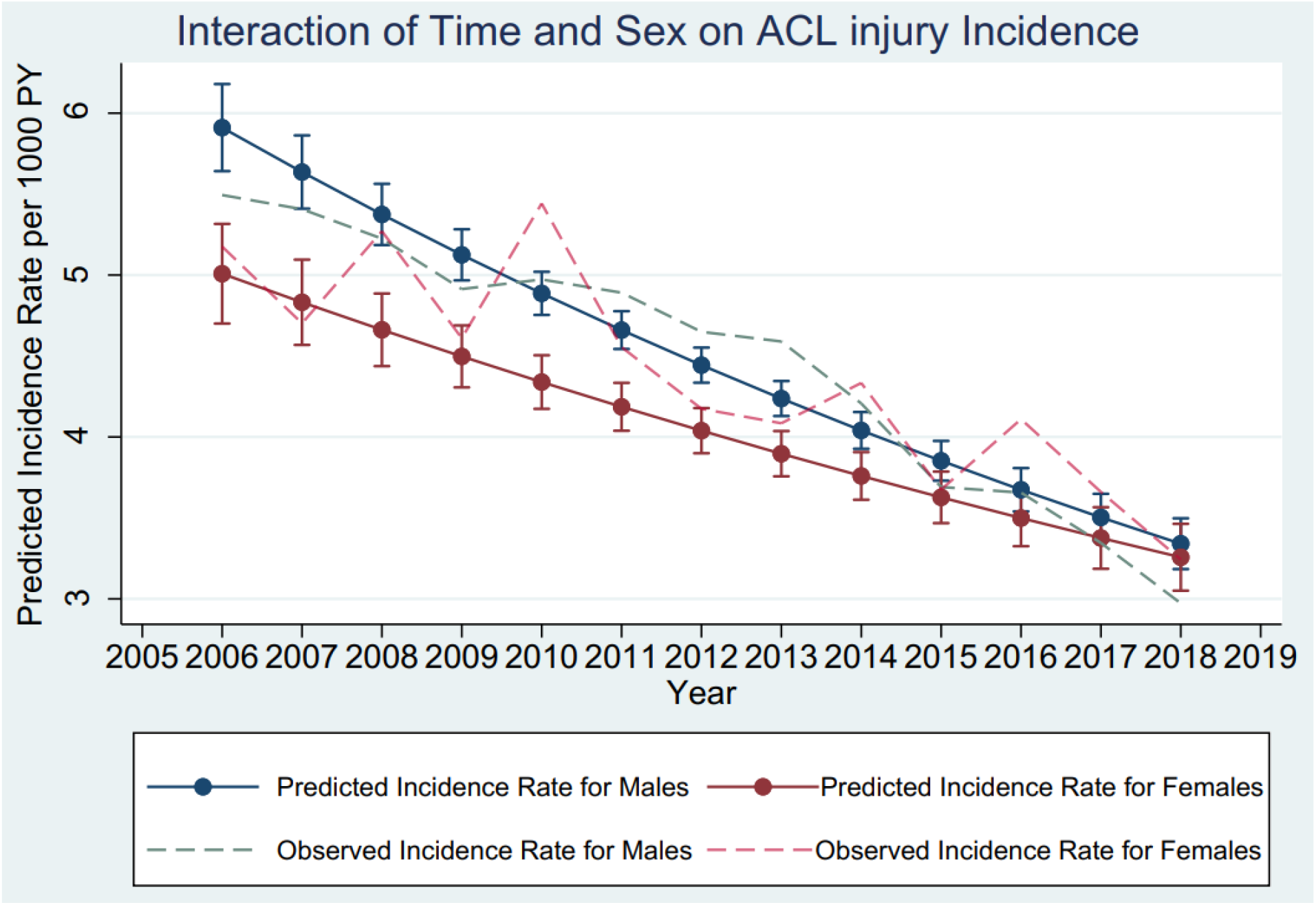
Predicted and Observed ACL Injury Incidence among Males and Females in the US Armed Forces, 2006-2018.

## DISCUSSION

The primary finding was that the study period displayed an overall statistically significant decrease in rates of ACL injury over time regardless of sex, rank, or branch of service. The interaction effect of time with sex indicated that the decrease in ACL injury incidence declined at a steeper rate in males as compared to females. This study found that the risk of ACL injury by sex was also modified by rank (officer versus enlisted). The risk of ACL injury among officers and enlisted personnel varied depending on occupation.

### Injury Rates Over Time

The significant decrease in incidence rate of ACL injury likely represents a real trend with a potential contributing factor due to changes in the operational tempo in the U.S. Armed Forces over the study epoch. This could be plausibly explained by a decreased exposure to hazards with the decline of operational demands. The higher rates of ACL injury at the beginning of this study in 2006 represents a time of increased military operations with a high frequency of multiple deployements involved in OIF and OEF with increased exposure to hazards that can lead to injury. Activity for these two campaigns peaked in 2008,^7^ and the campaigns concluded for OIF in 2011 and OEF in 2014.^33^ After controlling for sex and rank, all branches of service demonstrated a decrease in the incidence rate of ACL injury over time. The U.S. Army demonstrated statistically significant higher rates as compared to the U.S. Navy and Air Force.

Changes in coding may explain a small portion of this change as the ICD-10 coding transition was mandated to occur on 1 October 2015. However, even with this transition, the decline in ACL injury rates was already evident before this timepoint. While there should be a direct mapping of the codes used in this study, it is unclear how the capture of injury data for this study may have been affected.

### Sex and Time Interaction

As a group, males of the US Armed Forces had higher rates of ACL injury throughout the study epoch. Their rate of injury also declined faster over time from 2006 to 2018 as compared to females, on average, regardless of rank and branch of service as shown in Figure 5. Of note, the increase in incidence seen by female officers in the U.S. Marines during 2010 and 2016 may potentially coincide with the changes in physical fitness standards for the Combat Fitness Test in the USMC.^26^ There is a plausible influence that the changes in standards may have resulted in over engagement in risk-taking activity to meet these standards.

### Sex and Rank Interaction

Enlisted members, on average, had higher rates of ACL injury, but these relationships are more complex when stratifying by sex. The risk of ACL injury by sex demonstrated an effect modification depending on rank. In officers, female military members had a statistically significant higher risk compared to male officers regardless of occupation. This is contrasted by female enlisted members who were at a statistically lower risk compared to enlisted males regardless of occupation. The prior population-based study by Owens and colleagues did not stratify by rank^27^ which potentially masked risk differences between males and females in this study.

The relationship between sex and rank is noteworthy when compared to what is typically reported in the athlete population. When the risk of ACL injury is compared relative to the numbers of male and female athletes, the incidence of ACL injury is higher in females.^6^ A recent systematic review and meta-analysis reported that ACL injury was sustained by female athletes at 3 times the rate of male athletes in the same contact sports and were over 5 times greater for “fixed-object high-impact” sports such as obstacle course tests and races.^23^ Furthermore, when highly active groups of military members were studied, documented cases of higher incidence of ACL injury were reported in females.^18^ During the time period of this study, enlisted personnel were approximately 7.4 to 7.5 years younger than officers^1^ and were considerably closer in age to the athletes referenced in these studies. Therefore, it was surprising to find that the relative risk of ACL injury was lower in female enlisted personnel compared to their male counterparts regardless of occupation for this younger group within the military. This finding in combination with the trend reversal seen in officers challenges the current paradigm that younger, physically active females are the group at higher risk for ACL injury.

This provides an opportunity for hypothesis generation; elucidating what key factors within military members differ from competitive athletes may provide insight into why ACL injury trends differ by sex depending on rank. An important difference between these groups is that men and women train side by side in the military compared to training for most competitive sports that is clearly stratified by sex. While service members are required to meet physical fitness testing standards that are both age and sex specific, this does not hold true for occupational standards. Furthermore, it is likely that the proportions of women in previously all male occupations has yet to reach its peak even though women have been incorporated into all military occupations since 2016.^11^ This may be an influential factor on the differences in the rate of decrease in incidence seen by males and females over time. When higher proportions of women enter these fields, it is plausible that this could change the relationship of ACL injury and sex and how it relates to rank structure and military occupation. Finally, it is possible that young, enlisted males are participating in other activities outside of their military occupation that are increasing the risk for ACL injury in this group compared to their female counterparts.

### Occupation and Rank

Amongst officers and enlisted, the risk of ACL injury varied depending on occupation. Exposures to hazards may contribute to the occupational differences observed. This study found that enlisted occupations where vehicles were primarily employed or were more sedentary in nature compared to infantry had lower risk of ACL injury. Aviation, maintenance, administration, intelligence, communication, and maritime/naval specialties were at a statistically significant lower risk compared to infantry. Occupations where the knee is loaded on uneven surfaces are a known predisposing factor for ACL injury.^9^ Infantry members have higher exposures to hazards that can lead to ACL injury with increased outdoor activities such as rucking, maneuvering, and training over variable terrain in comparison to other occupations. Similarly, in the officer community, aviation and services officers had a statistically significant lower risk of ACL injury compared to ground and naval gunfire officers.

Special Operations Forces and Artillery/Gunnery occupations were at a statistically higher risk of ACL injury compared to infantry. The highest risk of ACL injury by occupation found in this study was in the Special Operations Forces; this may be explained by the increased intensity and frequency of tactical training with a correspondingly high level of musculoskeletal injury that is known to occur in this community.^2^ Occupation-specific training and physical activity levels alone are likely not the only important factors driving ACL injury. Risk-taking behaviors that are culturally influenced and vary by occupational communities are likely contributing factors as well.

### Social Determinants

Social determinants that affect health care utilization may play a role in the findings of this study. ACL injury is not typically self-limiting, and the billed medical encounters used to generate the data in this study are more likely to represent the true incidence. There is evidence that underreported injuries found in the US Army are due to chronic, overuse injuries as compared to acute musculoskeletal injuries.^32^ Due to the severity of an acute ACL injury, bias due to health care utilization is not likely to have a large impact on the study results. However, even with ACL injury, barriers to seeking care by military members should be considered, especially in those deemed as “copers” who suffer an ACL injury yet are still able to perform functional tasks. Fear that future career opportunities may be negatively affected is a concern among military members that may result in underreporting of injuries.^32^ Additional reasons that may affect reporting of ACL injury by military members include the service member’s perception of the convenience and quality of medical care they will receive.^15^ The cultural environment of the military reinforces the desire to put aside pain associated with an injury to ensure that the mission is completed and to avoid the negative perceptions associated with injury.^15^ It is plausible that underreporting may disproportionately affect certain occupations more than others,^12^ and this may have had an influence in the results of this study.

### Clinical and Research Implications

The results of this study highlight important trends of ACL injury in regards to sex, occupation, rank, branch of service, and changes over time that require further investigation into the cause of these trends. Specific hazards and exposures associated with military occupations should be explored in order to mitigate the risks. This is especially critical in communities such as Special Operations Forces, where a relatively smaller number of highly trained service members must perform demanding physical tasks which are crucial to complete the mission. Surveillance of ACL injury should continue as the percentage of women in previously restricted combat roles grows. It is essential for policy makers to understand the salient risk factors for ACL injury among the military population as a whole and within subgroups of this population to guide appropriate proactive measures to prevent injury. As rehabilitation specialists across the military branches continue to be incorporated into patient-centered medical homes and are assigned to operational units, the effect on injury risk, rehabilitation, and return to duty rates should be investigated.

### Strengths and Limitations

The DMED allows for a population-based analysis which provides the best estimation of ACL injury incidence to be captured based on billed medical encounters. This permitted the calculation of risk for the known, important factor of sex associated with ACL injury and allowed for exploration of time trends and rank, branch of service, and military occupation which had been previously unexplored on a population-basis.

There are also important limitations associated with this study due to inherent constraints associated with DMED. While using initial encounters allowed for the calculation of incidence, this study is also limited in the ability to capture laterality of an injury, and a new injury on the contralateral side may not be counted as such. Important salient factors associated with ACL injury that have been identified in military members are unable to be measured with this database, to include factors such as medical history or body mass index.^34^

## CONCLUSION

There was a statistically significant decrease in the incidence of ACL injuries among members of the U.S. Armed Forces between the years 2006 to 2018 at an average rate of 0.18 cases per 1000 person-years or a 4.08% relative reduction each year. The rate of decrease over time differed between males and females, with males experiencing a higher rate of decline when averaging across the variables of rank and branch of service. The relationship of ACL injury incidence and sex was modified by rank as demonstrated by the significant increase in ACL injury risk experienced by female officers compared to their male colleagues (RR = 1.14), and the significant decrease in ACL injury risk experienced by enlisted females compared to their male colleagues (RR = 0.82). It is plausible that the physical demands and opportunity for exposure within specific military occupations in the enlisted and officer communities may play a role in the significant differences in ACL injury incidence among occupations reported in this study.

Despite the decline in incidence among members of the U.S. Armed Forces over time, the rates of ACL injury still remain higher than the general U.S. population. Sex, rank, branch of service, and military occupation have been found to be risk factors for ACL injury, however, more investigation is required to develop injury prevention programs within these groups.

## Data Availability

The data that support the findings of this study are available from the corresponding author, JJF, upon reasonable request.

## Acknowledgment

The authors would like to sincerely thank Dr. Charity G. Moore Patterson, PhD, MSPH for her statistical expertise and guidance in the coding and interpretation of the negative binomial regression analysis.

---

**Supplemental Table 1:** ACL injury counts, population at risk, and injury rates (per 1,000 person-years) by year for Male Officers

| **Counts** | 2006 | 2007 | 2008 | 2009 | 2010 | 2011 | 2012 | 2013 | 2014 | 2015 | 2016 | 2017 | 2018 | Total |
| --- | --- | --- | --- | --- | --- | --- | --- | --- | --- | --- | --- | --- | --- | --- |
| Army | 372 | 360 | 375 | 352 | 384 | 412 | 356 | 360 | 348 | 281 | 278 | 248 | 202 | 4,328 |
| Navy | 226 | 192 | 189 | 194 | 179 | 144 | 135 | 166 | 146 | 128 | 101 | 117 | 100 | 2,017 |
| Air Force | 307 | 300 | 257 | 209 | 193 | 224 | 206 | 188 | 189 | 155 | 148 | 145 | 152 | 2,673 |
| Marines | 82 | 84 | 84 | 74 | 101 | 85 | 80 | 79 | 62 | 58 | 66 | 62 | 48 | 965 |
| Total | 987 | 936 | 905 | 829 | 857 | 865 | 777 | 793 | 745 | 622 | 593 | 572 | 502 | 9,983 |
| **Population** |  |  |  |  |  |  |  |  |  |  |  |  |  |  |
| Army | 69,311 | 71,314 | 73,556 | 76,044 | 78,737 | 81,506 | 82,562 | 82,654 | 81,903 | 79,386 | 76,947 | 74,910 | 75,110 | 1,003,939 |
| Navy | 44,641 | 44,059 | 43,828 | 43,986 | 44,341 | 44,732 | 44,667 | 44,699 | 45,016 | 44,766 | 44,622 | 44,270 | 44,342 | 577,969 |
| Air Force | 58,175 | 55,391 | 53,124 | 53,237 | 53,485 | 52,821 | 52,042 | 51,692 | 50,635 | 48,324 | 47,944 | 46,467 | 46,317 | 669,654 |
| Marines | 17,900 | 18,280 | 18,869 | 19,626 | 20,223 | 20,886 | 20,727 | 20,296 | 19,777 | 19,436 | 19,307 | 19,406 | 19,713 | 254,446 |
| Total | 190,027 | 189,044 | 189,378 | 192,893 | 196,786 | 199,944 | 199,998 | 199,341 | 197,330 | 191,912 | 188,820 | 185,053 | 185,481 | 2,506,008 |
| **Rate** |  |  |  |  |  |  |  |  |  |  |  |  |  |  |
| Army | 5.4 | 5.0 | 5.1 | 4.6 | 4.9 | 5.1 | 4.3 | 4.4 | 4.2 | 3.5 | 3.6 | 3.3 | 2.7 | 4.3 |
| Navy | 5.1 | 4.4 | 4.3 | 4.4 | 4.0 | 3.2 | 3.0 | 3.7 | 3.2 | 2.9 | 2.3 | 2.6 | 2.3 | 3.5 |
| Air Force | 5.3 | 5.4 | 4.8 | 3.9 | 3.6 | 4.2 | 4.0 | 3.6 | 3.7 | 3.2 | 3.1 | 3.1 | 3.3 | 4.0 |
| Marines | 4.6 | 4.6 | 4.5 | 3.8 | 5.0 | 4.1 | 3.9 | 3.9 | 3.1 | 3.0 | 3.4 | 3.2 | 2.4 | 3.8 |
| Total | 5.2 | 5.0 | 4.8 | 4.3 | 4.4 | 4.3 | 3.9 | 4.0 | 3.8 | 3.2 | 3.1 | 3.1 | 2.7 | 4.0 |

**Supplemental Table 2:** ACL injury counts, population at risk, and injury rates (per 1,000 person-years) by year for Female Officers

| **Counts** | 2006 | 2007 | 2008 | 2009 | 2010 | 2011 | 2012 | 2013 | 2014 | 2015 | 2016 | 2017 | 2018 | Total |
| --- | --- | --- | --- | --- | --- | --- | --- | --- | --- | --- | --- | --- | --- | --- |
| Army | 78 | 60 | 72 | 65 | 74 | 87 | 75 | 70 | 76 | 76 | 65 | 52 | 63 | 913 |
| Navy | 29 | 37 | 37 | 46 | 28 | 44 | 34 | 32 | 35 | 24 | 27 | 36 | 35 | 444 |
| Air Force | 61 | 52 | 75 | 66 | 62 | 62 | 59 | 57 | 61 | 49 | 53 | 37 | 48 | 742 |
| Marines | 9 | 6 | 9 | 3 | 16 | 3 | 5 | 6 | 8 | 6 | 13 | 10 | 5 | 99 |
| Total | 177 | 155 | 193 | 180 | 180 | 196 | 173 | 165 | 180 | 155 | 158 | 135 | 151 | 2,198 |
| **Population** |  |  |  |  |  |  |  |  |  |  |  |  |  |  |
| Army | 12,563 | 12,858 | 13,418 | 14,189 | 14,876 | 15,498 | 15,883 | 16,059 | 16,158 | 15,921 | 15,666 | 15,492 | 15,859 | 194,440 |
| Navy | 7,671 | 7,612 | 7,704 | 7,901 | 8,141 | 8,427 | 8,625 | 8,869 | 9,137 | 9,328 | 9,639 | 9,919 | 10,262 | 113,235 |
| Air Force | 13,032 | 12,263 | 11,784 | 11,967 | 12,203 | 12,182 | 12,233 | 12,518 | 12,531 | 12,165 | 12,334 | 12,374 | 12,623 | 160,209 |
| Marines | 1,127 | 1,124 | 1,161 | 1,201 | 1,264 | 1,312 | 1,354 | 1,379 | 1,426 | 1,454 | 1,507 | 1,566 | 1,639 | 17,514 |
| Total | 34,393 | 33,857 | 34,068 | 35,258 | 36,483 | 37,419 | 38,096 | 38,825 | 39,253 | 38,867 | 39,145 | 39,351 | 40,383 | 485,398 |
| **Rate** |  |  |  |  |  |  |  |  |  |  |  |  |  |  |
| Army | 6.2 | 4.7 | 5.4 | 4.6 | 5.0 | 5.6 | 4.7 | 4.4 | 4.7 | 4.8 | 4.1 | 3.4 | 4.0 | 4.7 |
| Navy | 3.8 | 4.9 | 4.8 | 5.8 | 3.4 | 5.2 | 3.9 | 3.6 | 3.8 | 2.6 | 2.8 | 3.6 | 3.4 | 3.9 |
| Air Force | 4.7 | 4.2 | 6.4 | 5.5 | 5.1 | 5.1 | 4.8 | 4.6 | 4.9 | 4.0 | 4.3 | 3.0 | 3.8 | 4.6 |
| Marines | 8.0 | 5.3 | 7.7 | 2.5 | 12.7 | 2.3 | 3.7 | 4.3 | 5.6 | 4.1 | 8.6 | 6.4 | 3.1 | 5.7 |
| Total | 5.1 | 4.6 | 5.7 | 5.1 | 4.9 | 5.2 | 4.5 | 4.2 | 4.6 | 4.0 | 4.0 | 3.4 | 3.7 | 4.5 |

**Supplemental Table 3:** ACL injury counts, population at risk, and injury rates (per 1,000 person-years) by year for Enlisted Males

| **Counts** | 2006 | 2007 | 2008 | 2009 | 2010 | 2011 | 2012 | 2013 | 2014 | 2015 | 2016 | 2017 | 2018 | Total |
| --- | --- | --- | --- | --- | --- | --- | --- | --- | --- | --- | --- | --- | --- | --- |
| Army | 2,141 | 2,243 | 2,377 | 2,385 | 2,553 | 2,505 | 2,399 | 2,293 | 1,970 | 1,640 | 1,495 | 1,301 | 1,064 | 26,366 |
| Navy | 1,399 | 1,239 | 1,249 | 1,236 | 1,167 | 1,134 | 1,029 | 1,057 | 969 | 856 | 793 | 668 | 621 | 13,417 |
| Air Force | 1,229 | 1,254 | 1,200 | 1,099 | 1,070 | 1,132 | 1,105 | 1,041 | 950 | 807 | 907 | 705 | 699 | 13,198 |
| Marines | 979 | 1,032 | 952 | 1,032 | 995 | 1,011 | 991 | 854 | 749 | 631 | 626 | 585 | 540 | 10,977 |
| Total | 5,748 | 5,768 | 5,778 | 5,752 | 5,785 | 5,782 | 5,524 | 5,245 | 4,638 | 3,934 | 3,821 | 3,259 | 2,924 | 63,958 |
| **Population** |  |  |  |  |  |  |  |  |  |  |  |  |  |  |
| Army | 356,602 | 368,968 | 385,276 | 397,813 | 406,484 | 407,158 | 392,488 | 375,912 | 357,011 | 339,619 | 325,696 | 321,308 | 322,174 | 4,756,511 |
| Navy | 252,588 | 241,732 | 235,115 | 232,772 | 228,757 | 224,608 | 218,499 | 217,744 | 218,836 | 219,525 | 218,162 | 213,620 | 216,444 | 2,938,404 |
| Air Force | 219,775 | 212,992 | 207,765 | 211,088 | 213,972 | 214,062 | 214,789 | 215,002 | 209,281 | 200,777 | 202,960 | 207,575 | 209,625 | 2,739,662 |
| Marines | 149,900 | 152,982 | 162,916 | 170,257 | 169,126 | 166,945 | 163,060 | 160,424 | 155,635 | 150,625 | 149,883 | 149,024 | 149,423 | 2,050,201 |
| Total | 978,865 | 976,674 | 991,073 | 1,011,930 | 1,018,340 | 1,012,773 | 988,837 | 969,083 | 940,763 | 910,546 | 896,701 | 891,528 | 897,665 | 12,484,777 |
| **Rate** |  |  |  |  |  |  |  |  |  |  |  |  |  |  |
| Army | 6.0 | 6.1 | 6.2 | 6.0 | 6.3 | 6.2 | 6.1 | 6.1 | 5.5 | 4.8 | 4.6 | 4.0 | 3.3 | 5.5 |
| Navy | 5.5 | 5.1 | 5.3 | 5.3 | 5.1 | 5.0 | 4.7 | 4.9 | 4.4 | 3.9 | 3.6 | 3.1 | 2.9 | 4.6 |
| Air Force | 5.6 | 5.9 | 5.8 | 5.2 | 5.0 | 5.3 | 5.1 | 4.8 | 4.5 | 4.0 | 4.5 | 3.4 | 3.3 | 4.8 |
| Marines | 6.5 | 6.7 | 5.8 | 6.1 | 5.9 | 6.1 | 6.1 | 5.3 | 4.8 | 4.2 | 4.2 | 3.9 | 3.6 | 5.4 |
| Total | 5.9 | 5.9 | 5.8 | 5.7 | 5.7 | 5.7 | 5.6 | 5.4 | 4.9 | 4.3 | 4.3 | 3.7 | 3.3 | 5.1 |

**Supplemental Table 4:** ACL injury counts, population at risk, and injury rates (per 1,000 person-years) by year for Enlisted Females

| **Counts** | 2006 | 2007 | 2008 | 2009 | 2010 | 2011 | 2012 | 2013 | 2014 | 2015 | 2016 | 2017 | 2018 | Total |
| --- | --- | --- | --- | --- | --- | --- | --- | --- | --- | --- | --- | --- | --- | --- |
| Army | 317 | 284 | 300 | 311 | 291 | 335 | 291 | 277 | 235 | 221 | 210 | 172 | 162 | 3,406 |
| Navy | 157 | 155 | 171 | 174 | 120 | 147 | 160 | 151 | 148 | 152 | 149 | 140 | 129 | 1,953 |
| Air Force | 218 | 227 | 184 | 209 | 190 | 157 | 164 | 174 | 159 | 148 | 127 | 150 | 145 | 2,252 |
| Marines | 55 | 57 | 55 | 59 | 74 | 78 | 54 | 52 | 63 | 44 | 45 | 54 | 49 | 739 |
| Total | 747 | 723 | 710 | 753 | 675 | 717 | 669 | 654 | 605 | 565 | 531 | 516 | 485 | 8,350 |
| **Population** |  |  |  |  |  |  |  |  |  |  |  |  |  |  |
| Army | 57,078 | 57,448 | 58,491 | 59,505 | 60,574 | 60,858 | 58,291 | 56,108 | 54,367 | 53,170 | 52,769 | 53,278 | 53,860 | 735,798 |
| Navy | 42,281 | 40,962 | 40,877 | 41,868 | 42,966 | 43,936 | 44,216 | 46,119 | 47,615 | 49,165 | 50,548 | 50,825 | 52,944 | 594,325 |
| Air Force | 55,031 | 53,268 | 51,798 | 52,144 | 51,625 | 50,689 | 50,062 | 49,662 | 48,046 | 46,445 | 47,724 | 49,930 | 51,819 | 658,244 |
| Marines | 9,955 | 10,320 | 10,954 | 11,622 | 12,120 | 12,419 | 12,484 | 12,710 | 12,816 | 12,703 | 13,165 | 13,716 | 14,136 | 159,121 |
| Total | 164,345 | 161,998 | 162,120 | 165,140 | 167,286 | 167,902 | 165,054 | 164,600 | 162,844 | 161,484 | 164,207 | 167,749 | 172,759 | 2,147,488 |
| **Rate** |  |  |  |  |  |  |  |  |  |  |  |  |  |  |
| Army | 5.6 | 4.9 | 5.1 | 5.2 | 4.8 | 5.5 | 5.0 | 4.9 | 4.3 | 4.2 | 4.0 | 3.2 | 3.0 | 4.6 |
| Navy | 3.7 | 3.8 | 4.2 | 4.2 | 2.8 | 3.3 | 3.6 | 3.3 | 3.1 | 3.1 | 2.9 | 2.8 | 2.4 | 3.3 |
| Air Force | 4.0 | 4.3 | 3.6 | 4.0 | 3.7 | 3.1 | 3.3 | 3.5 | 3.3 | 3.2 | 2.7 | 3.0 | 2.8 | 3.4 |
| Marines | 5.5 | 5.5 | 5.0 | 5.1 | 6.1 | 6.3 | 4.3 | 4.1 | 4.9 | 3.5 | 3.4 | 3.9 | 3.5 | 4.6 |
| Total | 4.5 | 4.5 | 4.4 | 4.6 | 4.0 | 4.3 | 4.1 | 4.0 | 3.7 | 3.5 | 3.2 | 3.1 | 2.8 | 3.9 |

